# COVID-19 vaccine information, misinformation, and vaccine uptake in Malawi

**DOI:** 10.1101/2023.07.14.23292688

**Authors:** John Songo, Hannah S. Whitehead, Khumbo Phiri, Pericles Kalande, Eric Lungu, Sam Phiri, Joep J. van Oosterhout, Agnes Moses, Risa M. Hoffman, Corrina Moucheraud

## Abstract

**Background:** COVID-19 vaccine information – including source, content, and tone – may be an important determinant of vaccination, but this dynamic is not well-understood in low-income countries where COVID-19 vaccine uptake remains low. We assessed the COVID-19 vaccine information environment in Malawi, and its correlation with vaccine uptake.

**Methods:** A survey was administered among 895 adult (≥18 years) clients at 32 Malawian health facilities in mid-2022. Respondents reported their COVID-19 vaccination history, exposure to information about the COVID-19 vaccine from different sources and its tone (positive, negative, or neutral/factual), and whether they had heard of and believed in ten COVID-19 and COVID-19 vaccine conspiracy theories. We described the COVID-19 vaccine information environment in Malawi and used logistic regression analyses to assess the association of exposure to information sources and conspiracy theories with uptake of the COVID-19 vaccine.

**Results:** Respondents had received information about the COVID-19 vaccine most commonly from friends and neighbors, healthcare workers, and radio (each reported by >90%). Men, urban residents, and respondents with a higher education level were exposed to more COVID-19 vaccine information sources. COVID-19 vaccine uptake was positively associated with exposure to a greater number of COVID-19 vaccine information sources (aOR 1.09, 95% CI 1.03-1.15), and more positive information (aOR 4.33, 95% CI 2.17-8.64) – and was negatively associated with believing COVID-19 vaccine conspiracy theories to be true (OR 0.76, 95% CI 0.68-0.87).

**Conclusions:** Malawian adults were exposed to a variety of COVID-19 vaccine information sources, with less access to information among women, rural residents, and people with lower educational attainment. Exposure to misinformation was common, though infrequently believed. Vaccination was associated with exposure to high number of COVID-19 vaccine information sources, exposure to positive vaccine information and endorsing fewer conspiracy theories. Vaccination programs should disseminate communication with positive messaging, through multiple information sources, prioritizing the less exposed groups we identified.

## Background

During public health crises, information can shape individuals’ knowledge of and/or attitudes about health interventions, and ultimately inform their behaviors (1,2). The COVID-19 pandemic, which occurred amidst widespread technology use and social media connectedness, was deemed an “infodemic” by the World Health Organization – i.e., it was characterized by an “overabundance of information,” both accurate and misleading, about the virus, its origins, and control measures, including vaccines (3). Misinformation about vaccines – i.e., incorrect or misleading information – can contribute to vaccine hesitancy (4). The spread of vaccination misinformation accelerated during the COVID-19 pandemic (5) and has been linked to decreased COVID-19 vaccination intent and behavior (6,7).

Two years into widespread availability of COVID-19 vaccines, most people in low-income countries have still not been vaccinated (8); and the COVID-19 information and misinformation landscape in these settings – and its association with vaccine uptake – remains poorly understood (9). In Malawi, a low-income country in southeastern Africa that has recorded nearly 90,000 cumulative confirmed COVID-19 cases and close to 3,000 deaths (10), only approximately 1 in 4 people have received a COVID-19 vaccination (11). Misconceptions and conspiracy theories about the COVID-19 vaccine have been documented in Malawi and throughout the region (12–14).

Globally, much of the literature on the COVID-19 “infodemic” focuses on the dissemination of (mis)information online and via social media (15,16), but it is not well-understood whether the internet is a major source of COVID-19 information in low-income countries. In countries like Malawi, access to technology is rapidly changing and there is a need to understand what this means for the spread of information during public health crises like the COVID-19 pandemic. Understanding what information people are exposed to, the sources and tone of this information, and how this information influences behaviors may help to inform vaccination promotion strategies in settings with low vaccine uptake. To this end, we conducted a survey to describe the information environment about COVID-19 and the COVID-19 vaccine in Malawi, and assessed how individuals’ exposure to information (from different sources, of differing tone) is correlated with vaccine uptake.

## METHODS

### Study design, site & participant selection

We conducted a cross-sectional survey of adults presenting at 32 purposefully-selected health facilities supported by Partners In Hope (PIH), a Malawian non-governmental organization that assists with implementation of the National HIV program across Malawi. Sites were selected to represent public and faith-based health facilities in urban, peri-urban, and rural areas across all three of Malawi’s regions (Northern, Central, and Southern). Eligible clients were at least 18 years old and visited a selected facility for care at an outpatient department (OPD), antiretroviral therapy (ART), or non-communicable disease (NCD) clinic during a time when the COVID-19 vaccine was available to the general public in Malawi. We used systematic random sampling (every second individual in queue to see a provider was approached) to invite individuals to participate in the survey.

### Survey Domains and Variable Definitions

The survey included questions asking whether respondents had been exposed to COVID-19 vaccine information from different sources, and if so, whether the information was positive, negative, or neutral/factual in tone. Respondents were also asked whether they had heard of ten common conspiracy theories about COVID-19 and the COVID-19 vaccine and if they believed them to be true; these 10 pieces of misinformation were selected by study team members as relevant to and circulating in Malawi from among those included in two prior COVID-19 surveys done in the UK and USA (17,18). We asked respondents about their experiences with COVID-19 and with COVID-19 vaccines, including vaccination details for those vaccinated (number of doses received, location, manufacturer, timing).

The survey instrument was developed in English and translated to Chichewa, the local language; it was then reviewed by bilingual (English/Chichewa) study team members to ensure clarity and meaning. *Exposure to COVID-19 vaccine information* was assessed by asking respondents if they had heard any information about the COVID-19 vaccine from thirteen different potential sources of information, spanning traditional media (newspaper; radio; television), health system and the government (health care provider; government/Ministry of Health), community figures (church or religious leaders; traditional medicine practitioner), personal relations (family member; friend; neighbor/community member), and social media (posts on Facebook, Instagram or Twitter from a known person; posts from an organization, company, or someone not known personally; conversations/groups in a messaging app such as WhatsApp or Facebook Messenger). In analysis, “friend” and “neighbor/community member” were combined and considered as a single information source. Exposure to information was quantified as the *count* of the number of different information sources reported, as well as the *proportion* of respondents who had heard information from a source.

*Tone of COVID-19 vaccine information* was measured as follows: for each source that a respondent reported hearing COVID-19 vaccine information from, they were asked whether they felt the information was overall positive, negative, neutral/factual, a mix of positive and negative, or that they did not know the tone of the information. In analysis, these responses were combined into three categories: positive or neutral, mixed tone or unknown, and negative. To quantify tone of information, for each respondent, we calculated the *proportion* of the information sources they reported exposure to that were positive or negative in tone (i.e., number of positive information sources divided by the total number of information sources exposed to), as well as *counts* of the number of positive or negative information sources. Respondents were also asked who they *trusted the most* to give them advice regarding COVID-19 vaccine.

*Vaccine uptake* was defined as having received any (1 or more) doses of any manufacturer’s COVID-19 vaccine (Johnson & Johnson, AstraZeneca or Pfizer).

*Exposure to COVID-19 misinformation* was characterized by asking whether the respondents had heard 10 common conspiracy theories about COVID-19 and the COVID-19 vaccine (17,18) and quantified as the *count* of heard conspiracy theories. For each conspiracy theory that the respondent had heard of, they were asked whether they believed it to be true, false, or did not know whether it was true or false.

*Endorsement of COVID-19 misinformation* was quantified as both the *proportion* of heard conspiracies believed to be true (number of conspiracies believed to be true divided by the number of conspiracy theories heard), as well as the *count* of conspiracies believed to be true.

### Data Collection

The survey was administered face-to-face as an interviewer-administered survey, in a private area within the health facility by a trained research assistant. All respondents provided oral informed consent for anonymous data collection prior to commencing the survey, and all responses were recorded using the SurveyCTO mobile data collection platform on Android tablets. All respondents received 4000 Malawi kwacha (approximately US$5) as compensation for opportunity costs.

Data were collected from 19 May to 30 June, 2022. At the time of data collection, all adults 18 years of age or older were eligible to receive the COVID-19 vaccine in Malawi. Ethical approval for the study was obtained from the National Health Sciences Research Committee in Malawi (#2883) and the University of California Los Angeles Institutional Review Board (#22-000380).

### Data analysis

We described sample characteristics, exposure to, and tone of COVID-19 vaccine information, and exposure to and endorsement of COVID-19 misinformation. Exposure to COVID-19 conspiracy theories was also visualized with a heat map table, in which cells were color-coded ranging from bright green (smallest proportion of respondents had heard theory) to red (highest proportion of respondents had heard theory). Chi-square tests, t-tests, and multivariable logistic regression models were used to assess whether these variables were associated with various socio-demographic characteristics. We used multivariable logistic regression models to assess whether COVID-19 vaccine uptake was associated with exposure to and tone of COVID-19 vaccine information, or exposure to and endorsement of COVID-19 vaccine misinformation. All multivariable logistic models were adjusted for gender, residence (urban/rural), age, HIV status, and education. All analyses were conducted using Stata v17.

## RESULTS

A total of 944 individuals were approached for participation in this survey; 46 declined to be screened or to consent to participation, and 3 were found to be ineligible (<18 years of age) during screening. The remaining 895 individuals (94.8% of those approached) completed the survey and are included in this analysis. Nearly half (43%) of respondents had received at least one dose of a COVID-19 vaccine. Over half of the respondents were female (57%) and three-quarters (75%) were married (Table 1). The majority of the respondents were residing in rural areas (82%), were employed (68%) and identified as Christian (91%).

**Table 1:**
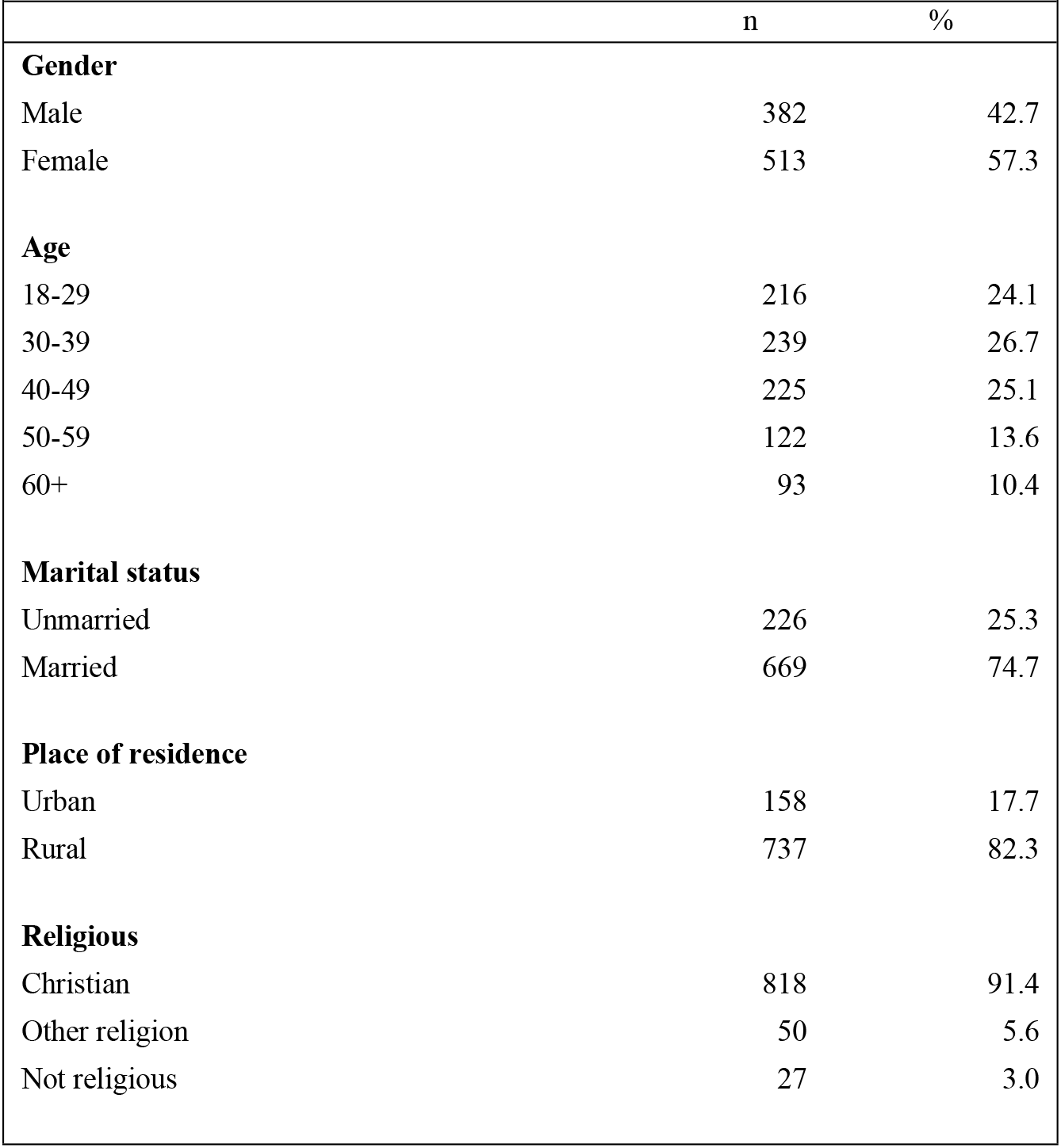
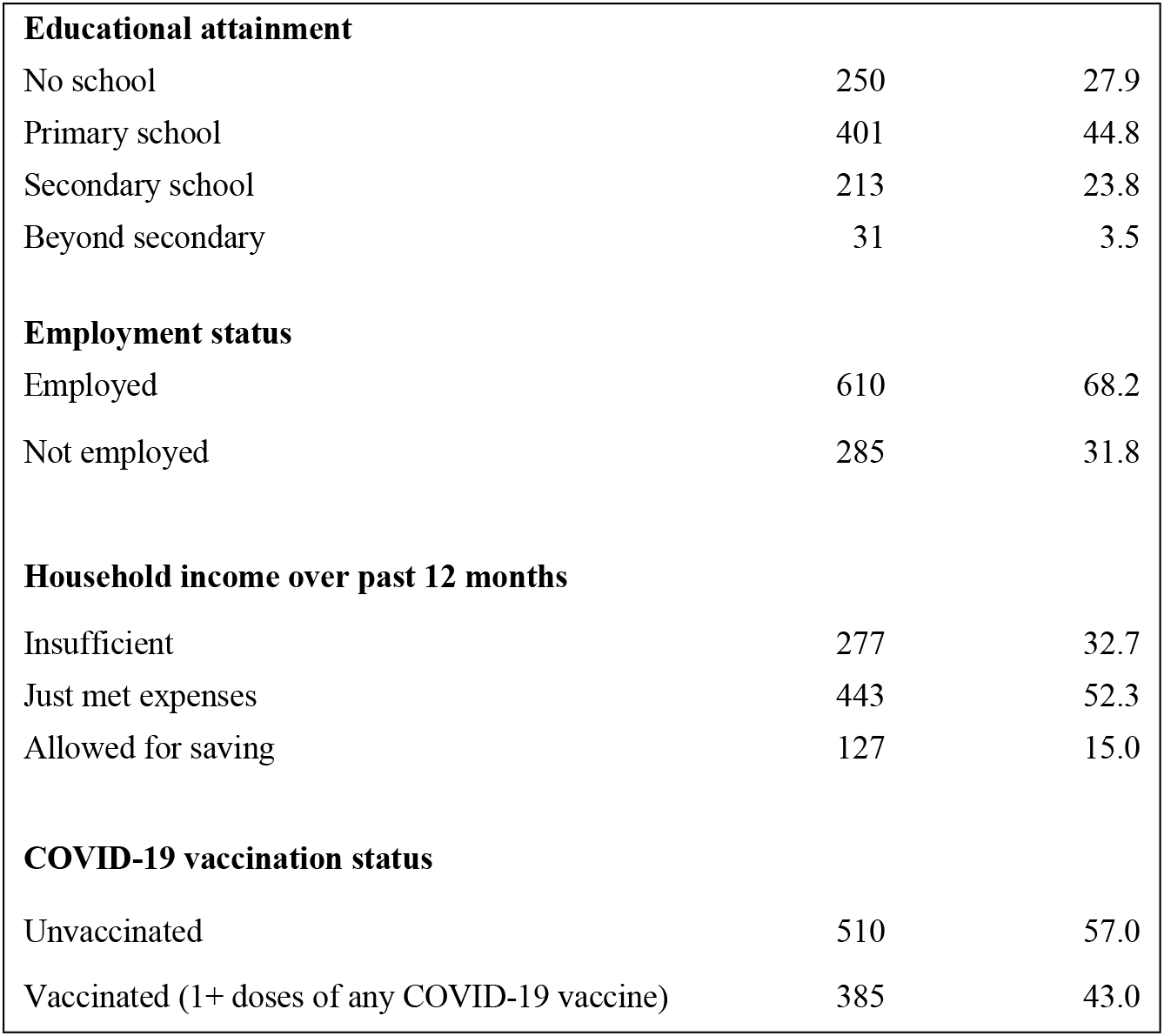
Sample characteristics (n=895)

| **Table 1: Sample characteristics (n=895)** |  |  |
| --- | --- | --- |
|  | n | % |
| **Gender** |  |  |
| Male | 382 | 42.7 |
| Female | 513 | 57.3 |
| **Age** |  |  |
| 18-29 | 216 | 24.1 |
| 30-39 | 239 | 26.7 |
| 40-49 | 225 | 25.1 |
| 50-59 | 122 | 13.6 |
| 60+ | 93 | 10.4 |
| **Marital status** |  |  |
| Unmarried | 226 | 25.3 |
| Married | 669 | 74.7 |
| **Place of residence** |  |  |
| Urban | 158 | 17.7 |
| Rural | 737 | 82.3 |
| **Religious** |  |  |
| Christian | 818 | 91.4 |
| Other religion | 50 | 5.6 |
| Not religious | 27 | 3.0 |
| **Educational attainment** |  |  |
| No school | 250 | 27.9 |
| Primary school | 401 | 44.8 |
| Secondary school | 213 | 23.8 |
| Beyond secondary | 31 | 3.5 |
| **Employment status** |  |  |
| Employed | 610 | 68.2 |
| Not employed | 285 | 31.8 |
| **Household income over past 12 months** |  |  |
| Insufficient | 277 | 32.7 |
| Just met expenses | 443 | 52.3 |
| Allowed for saving | 127 | 15.0 |
| **COVID-19 vaccination status** |  |  |
| Unvaccinated | 510 | 57.0 |
| Vaccinated (1+ doses of any COVID-19 vaccine) | 385 | 43.0 |

### Information about COVID-19 vaccines

#### Exposure to information about COVID-19 vaccine

All but one survey respondent had heard information about COVID-19 vaccines from at least one source. Respondents had been exposed to information from a median of 7 sources (IQR 6-9). The most common sources were friends and neighbors, health care workers, and radio – each reported by >90% of respondents (Figure 1). Receiving vaccine information from a traditional medicine practitioner was least common (3% of respondents), followed by social media, messaging apps, television and newspapers, each reported by 25-35% of respondents.

**Figure 1:**
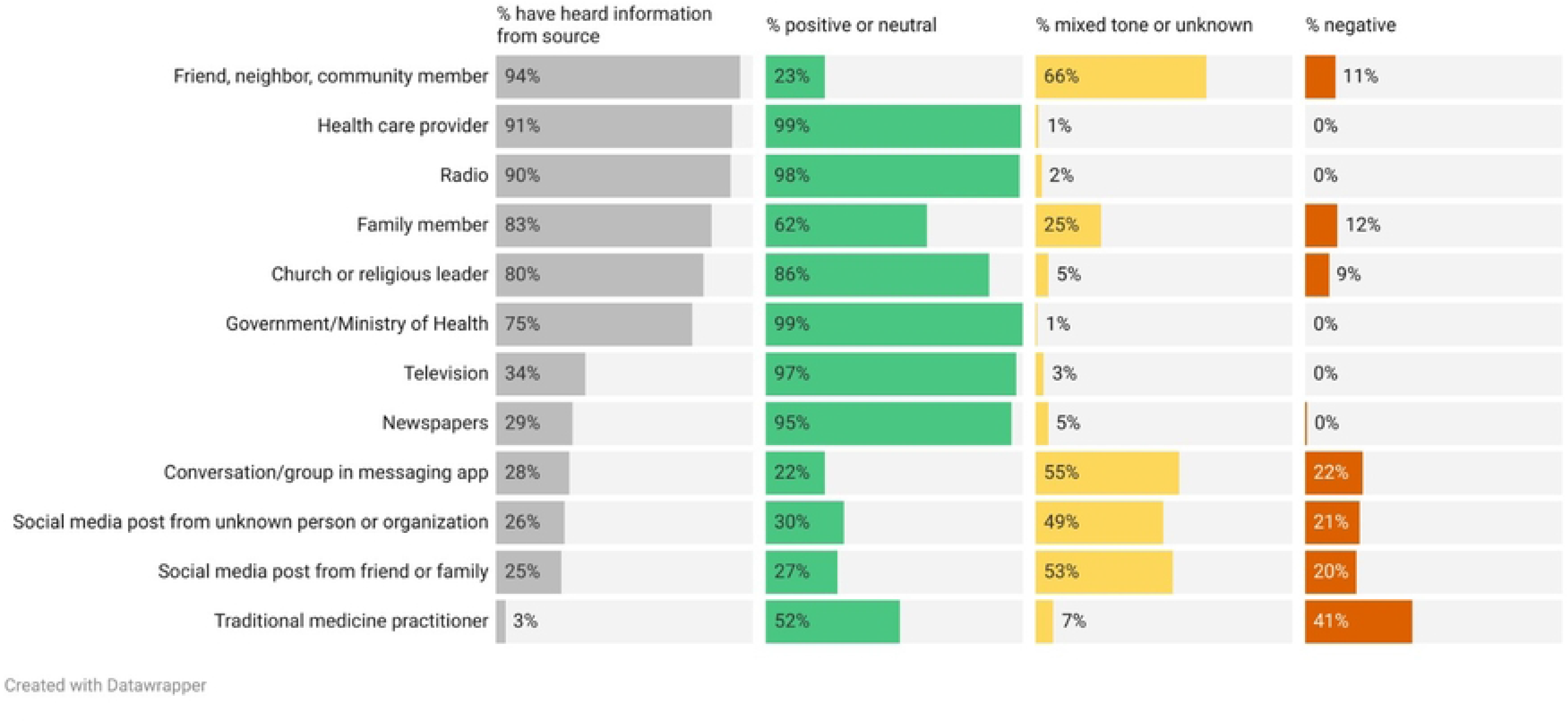
Exposure to information about COVID-19 vaccine, by source and tone.

Men, urban residents, and respondents with a higher education level were exposed to a higher mean number of COVID-19 vaccine information sources (men 8.2 vs. women 6.9; p<0.001; urban 8.6 vs. rural 7.2; p<0.001; secondary or higher education 9.3 vs. primary school or less 6.8; p<0.001). No differences were seen in the mean number of information sources by age, HIV status, or marital status.

Among specific information sources, men were significantly more likely than women to have heard COVID-19 information from nearly all sources, as were people with more educational attainment (Figure 2). Significantly less rural respondents reported exposure to information from traditional media (newspaper, television, radio) and social media than rural residents (OR 0.23, 95%CI: 0.079-0.609, p= 0.004)

**Figure 2:**
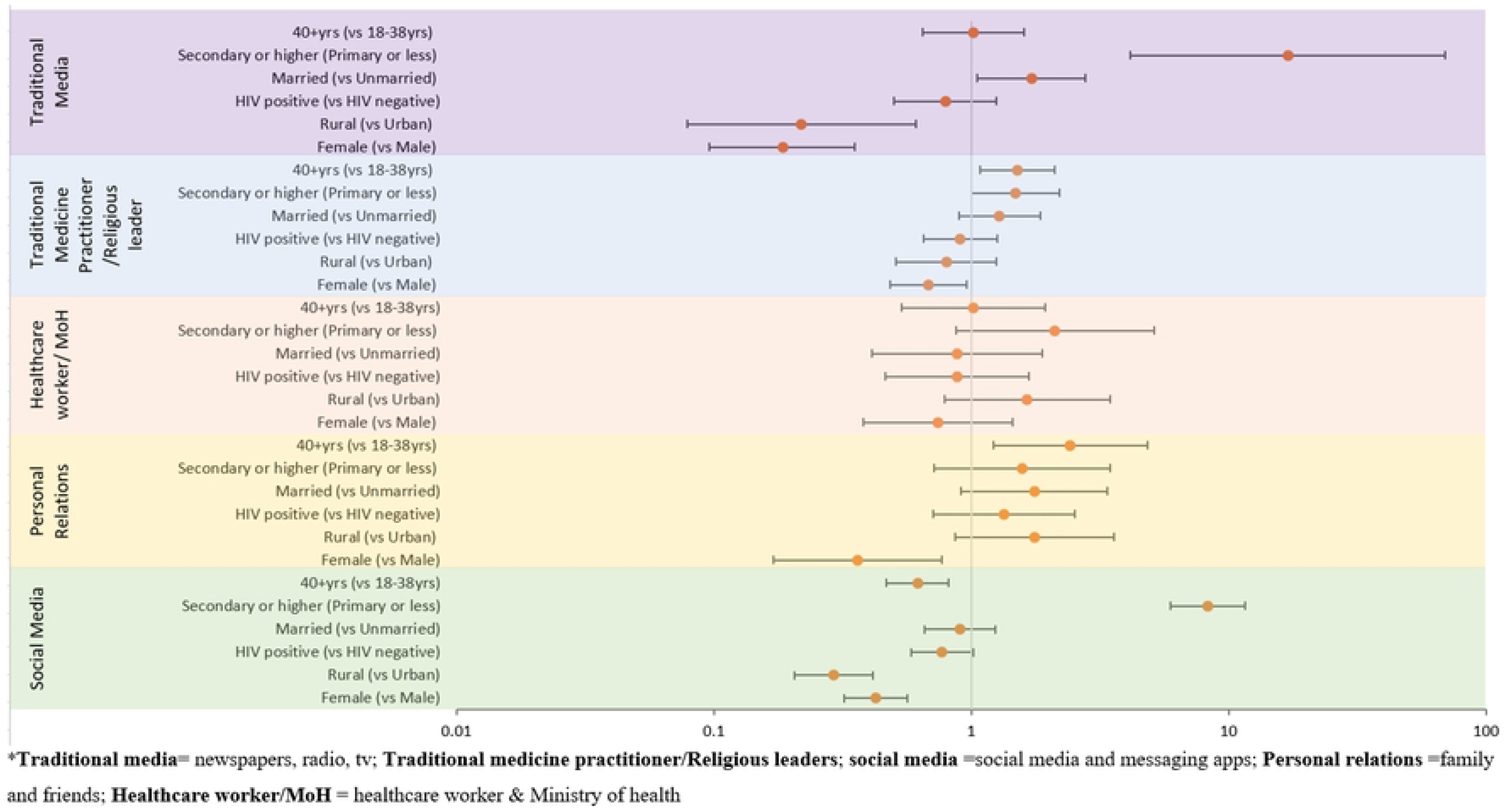
Odds of having received COVID-19 vaccine information from various sources, by respondent characteristics.

#### Tone of information about COVID-19 vaccine

Information about COVID-19 from health care workers, the government/Ministry of Health, religious leaders, friends and family, and traditional media (newspapers, television, radio) was largely (>80%) perceived as positive or neutral (Figure 1). Approximately one-fifth of people exposed to COVID-19 vaccine information on social media and messaging apps felt it was negative in tone and so did half of the people exposed to COVID-19 vaccine information from traditional medicine practitioners.

Women reported that a greater share of their information sources about COVID-19 vaccine was negative in tone: on average, women considered 10.2% of their information sources about COVID-19 vaccines negative in tone, versus 7.6% for men (p=0.015). Respondents with lower educational attainment reported that a lower share of their information sources was negative in tone (29% for those with a primary school education or less, versus 32% for those with secondary school education or higher, p=0.034). No differences were seen by HIV status, area of residence, or marital status.

#### Most trusted information sources

Respondents were asked who they trust most for advice regarding the COVID-19 vaccine. A healthcare provider was selected most often (69.4%), followed by a community health worker (22.1%); family, friends, religious leaders, and celebrities were selected infrequently (0.2-5% of respondents each). Unvaccinated individuals were twice as likely to most trust family, friends, or a religious leader for COVID-19 vaccine information as individuals who had been vaccinated (10.6% versus 5.2%, p=0.017). No significant differences in trusted information were seen by gender, HIV status, age, or education level.

#### Association of COVID-19 vaccine information with vaccine uptake

Having received information about COVID-19 vaccines from more sources was positively associated with COVID-19 vaccine uptake, with each additional source increasing vaccination probability by 9% (OR 1.09, 95% CI 1.03-1.15). Among subgroups, this relationship was found primarily among men (OR: 1.13, 95% CI: 1.04-1.23), respondents 40 years and older (OR: 1.15, 95% CI: 1.06-1.24), people living with HIV (OR: 1.17, 95% CI: 1.09-1.27), and those with a secondary school education or higher (OR: 1.15, 95% CI: 1.03-1.28) (Appendix 1). Adjusting for gender, residence (urban/rural), age, HIV status, and education, the positive association between information exposure (number of reported sources) and COVID-19 vaccination persisted (aOR 1.08, 95% CI 1.01-1.15) and the odds of vaccination was higher when respondents reported a greater proportion of COVID-19 information being positive in tone (aOR 4.33, 95% CI 2.17-8.64).

### Misinformation about COVID-19 vaccines

#### Exposure to misinformation about COVID-19 and COVID-19 vaccines

Respondents reported having heard a median of 5 of the 10 surveyed conspiracy theories. The most commonly heard were “The spread of COVID-19 is a deliberate attempt to reduce the size of the global population” (80.7% of respondents) and “The COVID-19 virus is a hoax” (74.1% of respondents) (Figure 3).

**Figure 3:**
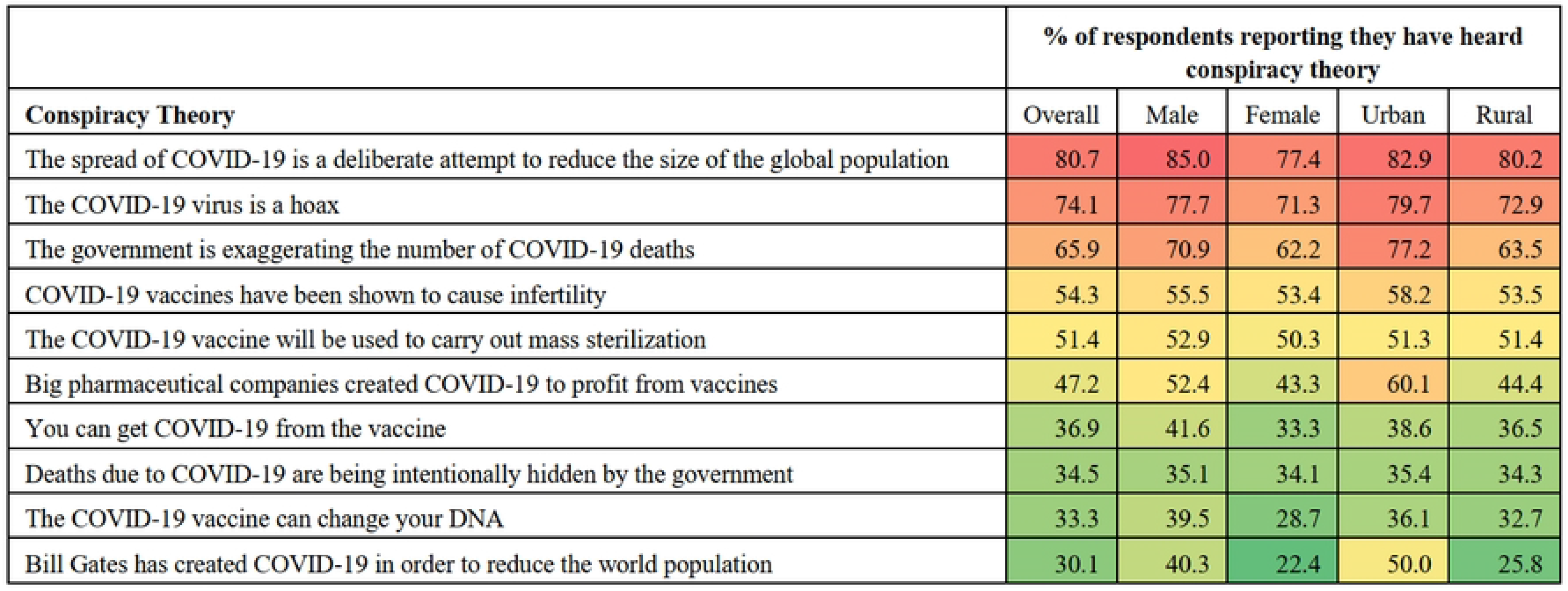
Heat map displaying the proportion of survey respondents (overall, by gender, and by place of residence) who reported hearing of 10 common COVID-19 conspiracies. Color scale spans from bright green (smallest proportion) to red (highest proportion).

Men, respondents with higher educational attainment, and urban respondents were exposed to a higher mean number of conspiracy theories: men 5.5 versus women 4.8 (p=0.0001); urban residents 5.7 versus rural residents 5.0 (p=0.002), and individuals with a secondary school education or higher heard 5.9 versus those with primary school or less 4.8 (p<0.001).

#### Endorsing misinformation about COVID-19 vaccines

Most respondents (72%) did not believe any of the 10 conspiracy theories to be true. Overall, respondents endorsed (believed to be true) a mean of 0.56 conspiracy theories; this increased to a mean of 1.96 among the 254 respondents who believed in at least one conspiracy theory. Younger age groups, urban respondents and respondents with higher education attainment endorsed more conspiracy theories: 18–39-year-olds endorsed 0.66 conspiracy theories versus 0.45 among 40+ year olds (p = 0.007), urban respondents endorsed 0.75 versus 0.51 among rural respondents (p=0.01), and individuals with a secondary school education or higher endorsed 0.76 versus 0.48 among those with primary school or less (p=0.001)

The most commonly endorsed myths were “The government is exaggerating the number of COVID-19 deaths” (22.7%), “Big pharmaceutical companies created COVID-19 to profit from vaccines” (17.3%), “The spread of COVID-19 is a deliberate attempt to reduce the size of the global population” (16.9%) and “Bill Gates has created COVID-19 in order to reduce the world population” (15.2%) (Table 2).

**Table 2:** Endorsement of 10 common conspiracy theories and association with COVID-19 vaccine uptake.

|  |  |  |  |  |
| --- | --- | --- | --- | --- |
| **Conspiracy theory** | **% Believe true, among those who have heard of the conspiracy theory** | **Odds of vaccination if believed theory to be true** |  |  |
|  |  | **OR** | **95% CI** | **p-value** |
| The government is exaggerating the number of COVID-19 deaths | 22.7 | 0.75 | 0.50-1.10 | 0.14 |
| Big pharmaceutical companies created COVID-19 to profit from vaccines | 17.3 | 0.44 | 0.26-0.75 | 0.00 |
| The spread of COVID-19 is a deliberate attempt to reduce the size of the global population | 16.9 | 0.33 | 0.21-0.52 | 0.00 |
| Bill Gates has created COVID-19 in order to reduce the world population | 15.2 | 0.4 | 0.20-0.80 | 0.01 |
| Deaths due to COVID-19 are being intentionally hidden by the government | 10.7 | 0.71 | 0.34-1.50 | 0.37 |
| The COVID-19 vaccine can change your DNA | 8.4 | 0.13 | 0.04-0.46 | 0.00 |
| You can get COVID-19 from the vaccine | 5.5 | 0.23 | 0.07-0.82 | 0.02 |
| The COVID-19 vaccine will be used to carry out mass sterilization | 4.1 | 0.27 | 0.09-0.82 | 0.01 |
| COVID-19 vaccines have been shown to cause infertility | 3.1 | 0.23 | 0.06-0.81 | 0.02 |
| The COVID-19 virus is a hoax | 2.9 | 0.22 | 0.06-0.75 | 0.01 |

#### Association with COVID-19 vaccine uptake

Exposure to a greater number of COVID-19 conspiracy theories was associated with increased odds of vaccine uptake (OR 1.10, 95% CI 1.05-1.16), while endorsing more conspiracy theories was associated with lower odds of COVID-19 vaccine uptake (OR 0.76, 95% CI 0.68-0.87). This association between conspiracy theory endorsement and vaccine uptake remained significant in univariate analyses among all considered subgroups (by gender, HIV status, age, and education) except urban residents (Appendix Table 2).

Respondents who believed that COVID-19 vaccines can change your DNA had the lowest odds of vaccination (OR 0.13, 95% CI 0.04-0.46), followed by those who believed that the COVID-19 virus is a hoax (OR 0.22, 95% CI 0.06-0.75) compared to those who believed these conspiracy theories to be false or didn’t know. Believing that the government is exaggerating the number of COVID-19 deaths and that deaths due to COVID-19 are being intentionally hidden by government were not associated with vaccine uptake.

## DISCUSSION

In a survey of adults presenting at health facilities in Malawi conducted while the COVID-19 vaccine was available to the general public, we found that people hear COVID-19 vaccine information from a variety of sources, and exposure to information – including the source, content, and tone – is associated with COVID-19 vaccine uptake. Exposure to more sources of information and to positive information were strongly associated with vaccine uptake. Endorsement of common COVID-19 conspiracy theories – though quite uncommon – was associated with decreased odds of vaccination.

Respondents heard COVID-19 vaccine information from a mean of 7 different sources. The 3 most common sources were peers, health care providers, and the radio. Information from health care providers, the Ministry of Health, traditional media, and church or religious leaders was mostly perceived as positive. Health care providers were reported as the most trusted source of COVID-19 vaccine information, as has been reported in other settings in sub-Saharan Africa (19,20). Information from peers (friends, neighbors, and community members) was largely mixed in tone, while information from social media, messaging apps, and traditional medicine practitioners was mostly perceived to be negative in tone. We found that the odds of vaccine uptake increased as the proportion of information sources that were positive in tone increased. Additionally, most people in our sample had heard several conspiracy theories about COVID-19, and endorsement of misinformation, while uncommon, was associated with decreased odds of vaccination. Negative vaccine information and misinformation have been reported in Malawi (21) and the harmful impacts on vaccine uptake have been well researched (5,15,16). Our findings underline the need for greater monitoring and action against vaccine information that is negative in tone as well as information that is intentionally misleading (15).

The degree of exposure to COVID-19 vaccine information varied across socio-demographic groups, with men, urban respondents, and individuals with more education tending to report exposure to a greater number of both vaccine information sources and conspiracy theories, likely reflecting greater literacy, internet/mobile penetration, and media access among these groups. Given the association between greater information exposure and vaccine uptake, these findings bolster previous studies that have found higher COVID-19 vaccine acceptance among males and individuals with more education (22). We also found greater endorsement of conspiracy theories among younger, less educated and urban respondents, possibly explaining findings from previous studies in sub-Saharan Africa that younger (23) and urban (24) respondents are more likely to be vaccine hesitant or believe in certain COVID-19 conspiracy theories, but conflicting with findings from Ghana that individuals with higher education are more likely to be vaccine hesitant(24). Our findings highlight demographic groups that may need to be reached with COVID-19 vaccine information, as well as where to target efforts at countering misinformation.

Our results point to important implications for programming to encourage COVID-19 vaccine uptake. Our finding that exposure to more information sources and information that is positive in tone is associated with increased vaccine uptake adds to an emerging literature on this topic (25,26) and is the first of its kind in Africa. This positive relationship has direct implications for communication strategies aimed at increasing vaccination. Campaigns should disseminate positive information through a variety of sources, and aim to reach less-exposed populations (such as women, rural residents, and those with less education) via multiple information sources, particularly those that have been shown to predominantly disseminate positive vaccine information (e.g., radio, television, or health care workers). Trust in healthcare workers should be leveraged by positioning them as spokespeople and vaccine “champions”, and by training them to share accurate, factual information about COVID-19 vaccines during one-on-one clinical encounters. While found to be low in our sample, endorsement of misinformation should be closely monitored, and vaccination programs may need to counter misinformation and conspiracy theories through culturally appropriate messaging targeted at the groups most susceptible to misinformation (15). Effective approaches would address and correct both negative and false information, particularly as spread by community members and via social media. More research is needed to identify effective and locally-relevant strategies for this in Malawi and similar contexts; one promising approach is a WhatsApp-based counselling intervention for countering COVID-19 fake news trialed in Nigeria (27). We note several limitations of this study. The study respondents were recruited at health facilities hence our sample is very likely to over-represent people with stronger trust in health care services, including COVID-19 vaccination. Vaccination status was ascertained based on self-report, which may have overestimated coverage and underestimated due to social desirability bias.

## Conclusion

This study represents one of the first in-depth explorations of the COVID-19 vaccine information environment and how it affects vaccine uptake in sub-Saharan Africa. Our findings provide important insights on how people’s vaccination behavior may be shaped by their local vaccine information sources, tone, and content. Investment is needed in targeted information interventions that leverage prominent and trusted sources of COVID-19 vaccine information, are designed to reach all socio-demographic groups, and combat misinformation.

## Data Availability

The data underlying the results presented in the study are available from (https://www.kaggle.com/datasets/johnpemphosongo/covid-19-vaccine-information-and-uptake-survey)

## Acknowledgements

We are grateful for the support of the participating health facilities and their management teams for allowing us to conduct surveys at the health facilities. We are also thankful to the research assistants who conducted the surveys.

## APPENDIX

Appendix 1: Association between hearing more information sources about COVID-19 vaccine, and COVID-19 vaccination, unadjusted odds ratios

| Subgroup | COVID-19 vaccination (OR) | 95% CI | p-value |
| --- | --- | --- | --- |
| Male | 1.13 | 1.04-1.23 | 0.004 |
| Female | 1.05 | 0.98-1.13 | 0.136 |
| Urban | 1.12 | 1.00-1.25 | 0.054 |
| Rural | 1.08 | 1.02-1.15 | 0.009 |
| HIV+ | 1.17 | 1.09-1.27 | 0.000 |
| HIV- | 1.01 | 0.94-1.09 | 0.724 |
| 18-39 years | 1.05 | 0.98-1.13 | 0.169 |
| 40+ years | 1.15 | 1.06-1.24 | 0.001 |
| Primary school or less | 1.06 | 0.99-1.13 | 0.111 |
| Secondary school or higher | 1.15 | 1.03-1.28 | 0.010 |

Appendix 2: Association between endorsing more conspiracy theories about COVID-19 and COVID-19 vaccine uptake

| Subgroup | OR for vax uptake | 95% CI | p-value |
| --- | --- | --- | --- |
| Male | 0.70 | 0.57-0.86 | 0.001 |
| Female | 0.82 | 0.68-0.98 | 0.026 |
| Urban | 0.86 | 0.67-1.09 | 0.209 |
| Rural | 0.72 | 0.61-0.85 | <0.001 |
| HIV+ | 0.77 | 0.64-0.94 | 0.009 |
| HIV- | 0.75 | 0.62-0.91 | 0.004 |
| 18-39 years | 0.82 | 0.68-0.99 | 0.034 |
| 40+ years | 0.74 | 0.61-0.91 | 0.004 |
| Primary school or less | 0.75 | 0.63-0.90 | 0.002 |
| Secondary school or higher | 0.75 | 0.60-0.92 | 0.006 |

## REFERENCE

1. Anwar A, Malik M, Raees V, Anwar A. Role of Mass Media and Public Health Communications in the COVID-19 Pandemic. Cureus [Internet]. 2020 Sep 14; Available from: https://www.cureus.com/articles/38293-role-of-mass-media-and-public-health-communications-in-the-covid-19-pandemic

2. Garfin DR, Silver RC, Holman EA. The novel coronavirus (COVID-2019) outbreak: Amplification of public health consequences by media exposure. Heal Psychol [Internet]. 2020 May;39(5):355–7. Available from: http://doi.apa.org/getdoi.cfm?doi=10.1037/hea0000875

3. World Health Organization. The COVID-19 infodemic [Internet]. Available from: https://www.who.int/health-topics/infodemic/the-covid-19-infodemic#tab=tab_1

4. Carrieri V, Madio L, Principe F. Vaccine hesitancy and (fake) news: Quasi-experimental evidence from Italy. Health Econ [Internet]. 2019 Nov 20;28(11):1377–82. Available from: https://onlinelibrary.wiley.com/doi/10.1002/hec.3937

5. Naeem S Bin, Bhatti R, Khan A. An exploration of how fake news is taking over social media and putting public health at risk. Heal Inf Libr J [Internet]. 2021 Jun 12;38(2):143–9. Available from: https://onlinelibrary.wiley.com/doi/10.1111/hir.12320

6. Roozenbeek J, Schneider CR, Dryhurst S, Kerr J, Freeman ALJ, Recchia G, et al. Susceptibility to misinformation about COVID-19 around the world. R Soc Open Sci [Internet]. 2020 Oct 14;7(10):201199. Available from: https://royalsocietypublishing.org/doi/10.1098/rsos.201199

7. Loomba S, de Figueiredo A, Piatek SJ, de Graaf K, Larson HJ. Measuring the impact of COVID-19 vaccine misinformation on vaccination intent in the UK and USA. Nat Hum Behav [Internet]. 2021 Feb 5;5(3):337–48. Available from: https://www.nature.com/articles/s41562-021-01056-1

8. United Nations Development Programme (UNDP). Global Dashboard for Vaccine Equity [Internet]. Available from: https://data.undp.org/vaccine-equity/

9. Safary E, Mtaita C. A qualitative exploration of perceptions of the COVID-19 vaccine in Malawi during the vaccine rollout phase. One Heal Implement Res [Internet]. 2022;2(2):79–87. Available from: https://ohirjournal.com/article/view/4965

10. Ministry of Health Malawi. COVID-19 National Information Dashboard [Internet]. Available from: https://covid19.health.gov.mw/

11. OurWorldInData.org. Coronavirus (COVID-19) Vaccinations [Internet]. Available from: https://ourworldindata.org/covid-vaccinations

12. Aron MB, Connolly E, Vrkljan K, Zaniku HR, Nyirongo R, Mailosi B, et al. Attitudes toward COVID-19 Vaccines among Patients with Complex Non-Communicable Disease and Their Caregivers in Rural Malawi. Vaccines [Internet]. 2022 May 17;10(5):792. Available from: https://www.mdpi.com/2076-393X/10/5/792

13. Wonodi C, Obi-Jeff C, Adewumi F, Keluo-Udeke SC, Gur-Arie R, Krubiner C, et al. Conspiracy theories and misinformation about COVID-19 in Nigeria: Implications for vaccine demand generation communications. Vaccine [Internet]. 2022;40(13):2114–21. Available from: https://www.sciencedirect.com/science/article/pii/S0264410X22001268

14. Dereje N, Tesfaye A, Tamene B, Alemeshet D, Abe H, Tesfa N, et al. COVID-19 vaccine hesitancy in Addis Ababa, Ethiopia: a mixed-method study. BMJ Open [Internet]. 2022 May 1;12(5):e052432. Available from: http://bmjopen.bmj.com/content/12/5/e052432.abstract

15. Gisondi MA, Barber R, Faust JS, Raja A, Strehlow MC, Westafer LM, et al. A Deadly Infodemic: Social Media and the Power of COVID-19 Misinformation. J Med Internet Res [Internet]. 2022 Feb 1;24(2):e35552. Available from: https://www.jmir.org/2022/2/e35552

16. Cinelli M, Quattrociocchi W, Galeazzi A, Valensise CM, Brugnoli E, Schmidt AL, et al. The COVID-19 social media infodemic. Sci Rep [Internet]. 2020 Oct 6;10(1):16598. Available from: https://www.nature.com/articles/s41598-020-73510-5

17. Freeman D, Loe BS, Chadwick A, Vaccari C, Waite F, Rosebrock L, et al. COVID-19 vaccine hesitancy in the UK: the Oxford coronavirus explanations, attitudes, and narratives survey (Oceans) II. Psychol Med [Internet]. 2022 Oct 11;52(14):3127–41. Available from: https://www.cambridge.org/core/product/identifier/S0033291720005188/type/journal_article

18. Hamel, Liz; Lopes, Luna; Ashley, Kirzinger; Sparks, Grace; Broddie M. KFF COVID-19 Vaccine Monitor: Media and Misinformation. KFF [Internet]. 2018; Available from: https://www.kff.org/coronavirus-covid-19/poll-finding/kff-covid-19-vaccine-monitor-media-and-misinformation/

19. Katoto PDMC, Parker S, Coulson N, Pillay N, Cooper S, Jaca A, et al. Predictors of COVID-19 Vaccine Hesitancy in South African Local Communities: The VaxScenes Study. Vaccines [Internet]. 2022 Feb 25;10(3):353. Available from: https://www.mdpi.com/2076-393X/10/3/353

20. Solís Arce JS, Warren SS, Meriggi NF, Scacco A, McMurry N, Voors M, et al. COVID-19 vaccine acceptance and hesitancy in low– and middle-income countries. Nat Med [Internet]. 2021 Aug 16;27(8):1385–94. Available from: https://www.nature.com/articles/s41591-021-01454-y

21. Chimatiro C, Hajison P, Jella C, Tshotetsi L, Mpachika-Mfipa F. Barriers affecting COVID-19 vaccination in Phalombe District, Malawi: A qualitative study. South African Med J [Internet]. 2023 Mar 8; Available from: https://samajournals.co.za/index.php/samj/article/view/842

22. Moola S, Gudi N, Nambiar D, Dumka N, Ahmed T, Sonawane IR, et al. A rapid review of evidence on the determinants of and strategies for COVID-19 vaccine acceptance in low– and middle-income countries. J Glob Health [Internet]. 2021 Nov 20;11:05027. Available from: http://jogh.org/documents/2021/jogh-11-05027.pdf

23. Ovenseri-Ogbomo G, Ishaya T, Osuagwu UL, Abu EK, Nwaeze O, Oloruntoba R, et al. Factors associated with the myth about 5G network during COVID-19 pandemic in sub-Saharan Africa. J Glob Heal Reports [Internet]. 2020 Nov 3;4. Available from: https://www.joghr.org/article/17606-factors-associated-with-the-myth-about-5g-network-during-covid-19-pandemic-in-sub-saharan-africa

24. Brackstone K, Atengble K, Head M, Boateng L. COVID-19 vaccine hesitancy trends in Ghana: a cross-sectional study exploring the roles of political allegiance, misinformation beliefs, and sociodemographic factors. Pan Afr Med J [Internet]. 2022;43. Available from: https://www.panafrican-med-journal.com/content/article/43/165/full

25. Kamal A, Hodson A, Pearce JM. A Rapid Systematic Review of Factors Influencing COVID-19 Vaccination Uptake in Minority Ethnic Groups in the UK. Vaccines [Internet]. 2021;9(10). Available from: https://www.mdpi.com/2076-393X/9/10/1121

26. Umakanthan S, Patil S, Subramaniam N, Sharma R. COVID-19 Vaccine Hesitancy and Resistance in India Explored through a Population-Based Longitudinal Survey. Vaccines [Internet]. 2021;9(10). Available from: https://www.mdpi.com/2076-393X/9/10/1064

27. Talabi FO, Ugbor IP, Talabi MJ, Ugwuoke JC, Oloyede D, Aiyesimoju AB, et al. Effect of a social media-based counselling intervention in countering fake news on COVID-19 vaccine in Nigeria. Health Promot Int [Internet]. 2022 Apr 29;37(2). Available from: https://academic.oup.com/heapro/article/doi/10.1093/heapro/daab140/6369161

